# Patient-Specific and Interpretable Deep Brain Stimulation Optimisation Using MRI and Clinical Review Data

**DOI:** 10.1101/2025.07.17.25331690

**Authors:** Apostolos Mikroulis, Andrej Lasica, Pavel Filip, Eduard Bakstein, Daniel Novak

## Abstract

**Background:** Optimisation of Deep Brain Stimulation (DBS) settings is a key aspect in achieving clinical efficacy in movement disorders, such as the Parkinson’s disease. Modern techniques attempt to solve the problem through data-intensive statistical and machine learning approaches, adding significant overhead to the existing clinical workflows. Here, we present an optimisation approach for DBS electrode contact and current selection, grounded in routinely collected MRI data, well-established tools (Lead-DBS) and, optionally, clinical review records.

**Methods:** The pipeline, packaged in a cross-platform tool, uses lead reconstruction data and simulation of volume of tissue activated to estimate the contacts in optimal position relative to the target structure, and suggest optimal stimulation current. The tool then allows further interactive user optimisation of the current settings. Existing electrode contact evaluations can be optionally included in the calculation process for further fine-tuning and adverse effect avoidance.

**Results:** Based on a sample of 177 implanted electrode reconstructions from 89 Parkinson’s disease patients, we demonstrate that DBS parameter setting by our algorithm is more effective in covering the target structure (Wilcoxon p<6e-12, Hedges g>0.34) and minimising electric field leakage to neighbouring regions (p<2e-15, g>0.84) compared to expert parameter settings.

**Conclusion:** The proposed automated method, for optimisation of the DBS electrode contact and current selection shows promising results and is readily applicable to existing clinical workflows. We demonstrate that the algorithmically selected contacts perform better than manual selections according to electric field calculations, allowing for a comparable clinical outcome without the iterative optimisation procedure.

## Introduction

Deep Brain Stimulation (DBS) is a treatment method for late-stage Parkinson’s disease (PD), aiming to minimise symptoms. [1] It involves precise electrical stimulation that aims to modulate aberrant neural activity. Despite its clinical success, DBS parameters have to be optimised for each patient, leading to complex, empirical adjustments over several clinical visits. As a result, traditional DBS optimisation protocols are time-intensive, costly and burdensome for the patients [2,3]

Modern optimisation approaches attempt to resolve these issues through machine learning methods, such as autoencoder-based feature extraction and probabilistic models.[4–6] These methods excel in predicting the optimal stimulation parameters based on complex datasets, including functional magnetic resonance imaging (fMRI) response maps. However, their accuracy often comes at the cost of explainability, which may limit their adoption in clinical settings, where recommendations require a clear reasoning. Additionally, these techniques frequently rely on large-scale, high-dimensional datasets, which can be resource-intensive and clinically impractical. More specifically, the most recent approaches require machine learning models to be pretrained or additional pre-computed data; this requirement can be a discriminant analysis model [4], a multilayer perceptron [7], a collection of pre-compiled statistical models [6], or fibre tract modelling and an additional, NEURON-based simulation step [8].

In this study, we propose an optimisation method for stimulation parameters (i.e. electrode contact and current selection) that can be directly optimised through electric field calculation and its overlap with the targeted structure, based entirely on anatomical data and analytic geometry. Our approach integrates magnetic resonance imaging (MRI) with a processing pipeline building upon well-documented DBS simulation tools to optimise DBS parameters in a manner that is minimally demanding in terms of data requirements.

Our processing pipeline utilises established processing tools: the Lead-DBS [9] reconstruction of the implant, to derive the geometry of individual electrode contacts and the target structure of the stimulation, and OSS-DBS [10] to perform fast, adjustable calculations of the reach of the electric field into the target structure. Clinical evaluations of the patient’s response to individual electrode contact and current selection during the initial clinical visit can be optionally integrated into the optimization procedure. Importantly, we prioritise a simple user interaction, only requiring a reconstruction of the implant (in Lead-DBS format) and clinical evaluation of the electrode contacts and currents, if available.

We are evaluating our method using retrospective clinical data targeting the Subthalamic Nucleus (STN), which is a common target with demonstrated involvement in generating Parkinson’s Disease symptoms.[11] This single-patient approach contrasts with prevailing machine learning-based methods by prioritising clinical utility, computational efficiency, and robust integration with existing clinical data collection and basic reconstruction workflows.

## Materials and Methods

### Patients and data

This study uses data from 104 patients with Parkinson’s disease who underwent bilateral implantation of deep brain stimulation electrodes (Medtronic 3389, Medtronic B33 series, or Abbott 6172), targeting the STN. In total, MRI data from 104 patients were processed, of which 89 also had corresponding clinical review information and at least one hemisphere with documented stimulation settings. This yielded 177 implantation instances meeting all inclusion criteria for direct comparison.

All participants provided written informed consent to participate in the study upon enrolment. The study was approved by Ethics Committee of the General University Hospital in Prague (case number 59/18) and conducted in alignment with the Declaration of Helsinki.The data were sourced retrospectively from records of the iTEMPO department of the Neurology clinic, General University Hospital in Prague). For this dataset the (average ± standard deviation) age at PD onset was 45 ± 8.6 (n=64), and the age at DBS surgery was 56.2 ± 8.8 (n=89). A discrepancy between the numbers of patients with a PD onset record compared to the DBS surgery record is noted, due to incomplete reporting for some of the patients (hospital transfers).

We incorporated clinical evaluations of electrode contacts, done during initial stimulation setup at a clinical visit after implantation. The records indicated that the clinical review was performed in four contact group configurations per hemisphere: (i) individually activated edge contacts, (ii) separately tested middle non-directional contacts, or two groups of three radially arranged directional contacts in the case of directional leads. These groups were stimulated across incremental current levels ranging from 0.5 mA to 4.0 mA, with assessments focused on clinical improvement in rigidity, akinesia, and tremor, as well as thresholds for adverse effects.

Statistical analyses were conducted using the SciPy and Pingouin libraries, with Friedman tests followed by Bonferroni-corrected Wilcoxon signed-rank tests for pairwise comparisons.

### MRI data processing

Pre-operative (3 T) and post-operative (1.5 T) MRI images (Nifti format) were imported into the Lead-DBS toolbox [9], for atlas co-registration (using the SPM method [12]), normalisation (using the ANTs method [13] with SyN nonlinear transform and mutual information metric), subcortical brainshift correction [14], and electrode reconstruction performed manually in Lead-DBS. All subsequent volume and coordinate processing was done after conversion back to the native patient space for each subject.

### Optimisation method

The proposed method, outlined below, consists of two steps: i) contact selection based on spatial configuration of the STN and the stimulation lead, and ii) current selection, based on modelling of the volume of tissue activated (VTA). In the second step, two variants of the method were evaluated: a) geometry-based method only, and b) a variant considering the clinical review with test stimulation. The method is available as a standalone python-based GUI tool (Figure S2).[15]

#### 1. Contact selection

##### Geometry score

To identify the optimal contacts, we calculated their spatial relationship to the centre of mass of the motor subregion of the STN. Two geometric features were considered for each contact: (i) the Euclidean distance to the motor STN centroid, and (ii) the rotation angle between the contact and the centroid, relative to the electrode axis (for directional contacts). As these metrics differ in scale and units, they were independently ranked from lowest to highest across all contacts. The ranks were then summed to yield a geometry-based score for each contact *s*_*geometry,C*_. For non-directional electrodes, and for edge contacts on directional electrodes where rotation is undefined, only distance was used; in such cases, a nominal angle of 90° was assigned to preserve consistency across contact types.

##### Clinical-review based score

The clinical review data only contained coarse contact groups (radially distributed directional contacts were evaluated as a single contact, making up a total of four coarse contacts in all types of electrodes). The clinical review thus evaluated four contact groups in all lead types.

The clinical evaluation data contained rigidity *s*_*rigidity,i,C*_, akinesia *s*_*akinesia,i,C*_, and tremor *s*_*tremor,i,C*_ scores, on a scale from zero (no symptoms) to six for each current setting at each contact group, along notes where stimulation adverse effects occurred. The total clinical score *s*_*i,C*_ for every current step *i* and coarse contact *C* was calculated from the rigidity, akinesia, and tremor scores:

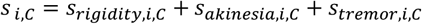

The higher-current positions where the adverse effects occurred were excluded from the evaluation. An improvement is denoted by a decreasing clinical score sum, compared to the clinical score sum at the previous current step.

The difference of the total clinical score was evaluated for successive current steps, *i* (up to the last adverse effect-free current step, *I*) and scaled by the number of current steps to prioritise faster improvements (with lower current). The cumulative sum of the scaled differences was calculated for every current step, and scaled by the initial total clinical score (current step *i* = 0, with no current applied):

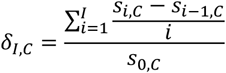

lower negative values of *δ*_*I,C*_ thus meaning greater improvement.

The best improvement for every contact was compared to the average of the improvements of all four contacts:

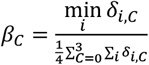

The clinical review scaling factor *κ* was calculated by scaling the improvements of each contact to add up to 1, and calculating their ratio over ¼ (which would be their value if all four contacts achieved the same non-zero improvement):

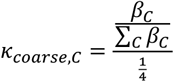

Since the clinical review performed in the clinical centre makes no distinction between the directional contacts (*c*) for every group of contacts *C*, the same value was used for all included directional contacts:

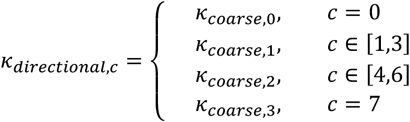

For the non-directional electrodes, the coarse evaluation contacts correspond to the physical contacts, so the same values were used:

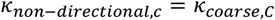

Depending on the electrode type (directional or non-directional), *κ*_*c*_ was set to *κ*_*directional,c*_ or *κ*_*non*−*directional,c*_, respectively. The scaling factor was applied as 1 + (1 − *κ*_*c*_) (to centre it around 1, with the improvement towards 0) for each geometry-based contact score *s*_*geometry,c*_,

##### Combined score

For each contact, a coefficient based on the centered clinical scaling factor *κ*_*c*_ was added to the geometry score *s*_*geometry,C*_ with 50% weight, according to:

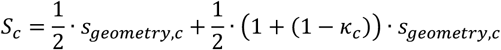

Based on the value of the combined score *S*_*c*_, the contacts with the two to three lowest scores were selected. The third contact was added for directional leads, only if (a) its score was tied with the second contact, or (b) if the score difference between the second and third was smaller than that between the first and second contacts and also smaller than the score of the top-ranked contact

For the non-directional electrodes, the selection was restricted to the two best contacts. The second contact was added if its score difference from the first contact was smaller than the first contact’s score.

#### 2. VTA modelling and overlap

The volume of activated tissue was calculated using OSS-DBS v2 [10], at an electric field threshold of 200 mV/mm [16]. The standalone OSS-DBS was preferred over the default Lead-DBS-integrated methods in this case since it allows more flexibility in iterative settings modification for contacts and currents, and operates in the patient-specific native space, which eliminates discrepancies introduced by co-registration. The VTA voxel size was decreased from 0.33 mm to 0.2 mm, and the point model lattice dimensions were increased from the default 71^3^ to 119^3^ voxels to increase the VTA resolution. The default solver iteration limit was doubled to 1000 steps, to avoid non-converging runs with the increased resolution. The default behaviour of OSS-DBS generating a “success” file was disabled to allow multithreading.

The coordinates of the electrode contacts (including subcortical refinement/brainshift corrections) were retrieved from the pre-processed Lead-DBS reconstruction files. The DISTAL atlas[17] was used, since it includes the histological labels necessary to calculate precise electric fields, as well as STN parcellation into the motor, associative and limbic subregions.Similarly, the coordinates of the STN subregions were retrieved from the pre-processed Lead-DBS native-space coordinates files. The VTAs were generated directly by OSS-DBS in Nifti format.

To calculate VTA overlap with STN subregions, the coordinate sets provided by Lead-DBS were converted into contiguous volumetric representations by filling the convex hull of each region. Both VTA and STN volumes were upsampled 1000-fold (10× in each dimension) using third-order spline interpolation to minimise voxel size–related discretisation errors. The VTAs generated by OSS-DBS were already oriented correctly, with only diagonal and translational components present in the affine transformation matrix. Overlap metrics were computed as (i) the proportion of STN subregion voxels intersecting the VTA (motor, limbic and associative), and (ii) the fraction of VTA voxels intersecting the motor STN region.

#### 3. Current selection

Next, the following current selection was performed for the set of contacts, pre-selected in the previous step, using VTA calculation and its overlap with STN subregions.

To estimate the stimulation current needed, an initial VTA was calculated for a 4 mA current (*I*_*initial*_), which typically covers a substantial portion of the motor STN subregion and represents a typical upper-bound from a clinical perspective. The vectors from the midpoint of the active contacts (selected in the previous, contact selection step) to the vertices of this VTA were compared to the vector from the same midpoint of the active contacts to the centre of mass of the motor STN subregion. The three vertex vectors with the smallest angular deviation from the centroid vector were averaged (*r*_*VTA,initial*_). The ratio of the centroid vector length (*r*_*centroid*_) to this averaged vector length was squared and applied as a scaling factor to the initial 4.0 mA:

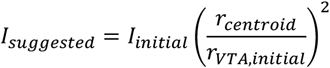

This approximates the current for a VTA reaching the centroid of the motor STN, assuming a constant conductivity in the distance range between the initial and the estimated VTA extents. The current was rounded to the closest 0.1 mA. For the single current selection, if the selected current was lower than 1 mA, 1 mA was selected, and clipped at 3.5 mA if it exceeded that value.

The final result consists of the contact and current selection, and the overlap of the VTA with motor and non-motor areas. An important output in judging the contact and current selection efficacy is the fraction of the VTA that is contained within the motor subregion. Additionally, we calculate the current-normalised values for these overlaps, to gauge the efficacy of the contact selection directly.

### A GUI tool for parameter setting

The aforementioned method is implemented as a Python-based GUI (Figure S2) app [15] and provides the recommended settings in CSV format. Additionally, to facilitate exploration of the space of possible parameter settings by the user, the VTAs for four more currents (0.8, 1.6, 2.4, 3.2 mA) can be optionally calculated (for a total of up to six current – VTA pairs). The resulting parameters including subregion overlaps are then interpolated using a cubic spline fit in the range from 0.5 to 4 mA and for the three subregions of the STN are presented in an interactive window. An additional assisting tool highlights a range of current values to optimise motor STN coverage against VTA outside the motor STN subregion, limbic STN coverage, and associative STN coverage, using a selectable minimum percentage of the corresponding harmonic mean range (over the entire current range).

### Output evaluation

The contact and current settings, suggested by the proposed method, were compared against the clinical DBS settings used by patients. As most patients were followed longitudinally, often with multiple adjustments over time, only the earliest complete set of stimulation parameters was analysed to minimise confounding effects from disease progression, electrode drift, or current increases. For patients programmed using voltage-controlled stimulation, current values were derived using the recorded electrode impedance. In cases involving interleaved stimulation, the effective current was calculated as a weighted average based on the duty cycle duration, pulse duration, and frequency of each setting.

The contact selection between the two methods (algorithm-based and clinical settings) was compared with a Jaccard index. For each implanted electrode, the cross-method Jaccard index was calculated from the ratio of coinciding contacts selected by both methods over the set of all selected contacts by either method. For directional contacts, all the subcontacts were considered for this calculation.

### Outcome prediction

Motor symptom improvement was estimated using a subset of patients (n = 50) who underwent UPDRS-III evaluation of DBS efficacy within three years post-surgery. Of these, a smaller group (n = 18) had their first UPDRS-III assessment conducted during the same visit in which DBS settings were first recorded (6–12 months postoperatively), while the remaining patients had delayed assessments occurring one to three years after surgery. For each patient, relative motor improvement *I* was calculated using paired UPDRS-III scores with stimulation turned on and off during the same visit:

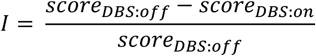

Two related variables were used to predict the improvement : (i) normalised motor-STN coverage as a measure of contact selection effectiveness, and (ii) current (mA) as a measure of stimulation strength affecting the size of the VTA.

A random forest regression model was trained using a maximum tree depth of seven, as performance gains plateaued beyond depth six and showed no further improvement at depths 8 or 9. To reduce variance and stabilise predictions, the model was configured with 5,000 estimators.

## Results

### Output overview

In total, the algorithm was evaluated on 177 implantations from 89 patients. The Figure 1 contains examples of the resulting stimulation settings, visualized using VTA approximation in Lead-DBS (Figure 1A-B), or by using directly the thresholded electric field data from OSS-DBS (Figure 1C-D).

**Figure 1.**
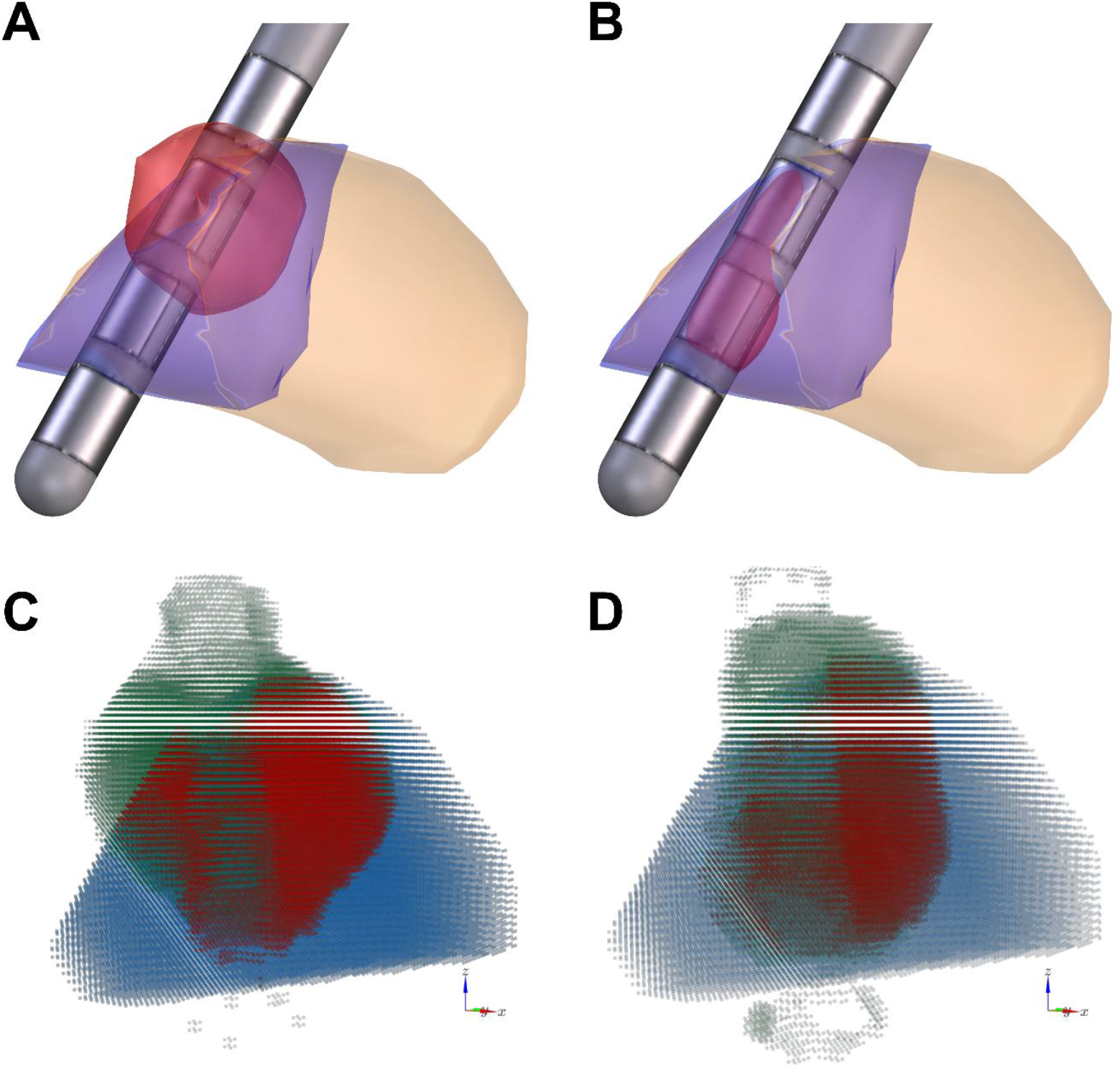
Visualisation of the VTA for clinically selected, patient settings and algorithm selected settings **(A-B)** Visualisation of the VTA approximation by Lead-DBS, using **(A)** patient settings and **(B)** algorithm-calculated settings (red: VTA approximation, light orange: STN, blue: motor subregion of the STN). **(C-D)** Visualisation of the VTA estimated by OSS-DBS, *with the same settings as (A-B)*, using **(C)** patient and **(D)** algorithm-calculated settings (green: VTA estimate, blue: motor STN subregion, red: overlap).

### Comparison with clinical settings

The contact selection was assessed in comparison to the clinical settings, in two different scenarios: (a) without any clinical information, and (b) using clinical review data (weighted at 50%). The contact selection overlapped in approximately 80% of the cases (Figure 2A-B). To quantify the overlaps, for each implant, a Jaccard index was calculated between the expert-selected contacts and the algorithm-selected contacts (expressed as the intersection of the two contact sets over their union). This quantification showed slightly increased partial overlaps with the expert contact selections when the clinical review information was used (Figure 2C).

**Figure 2.**
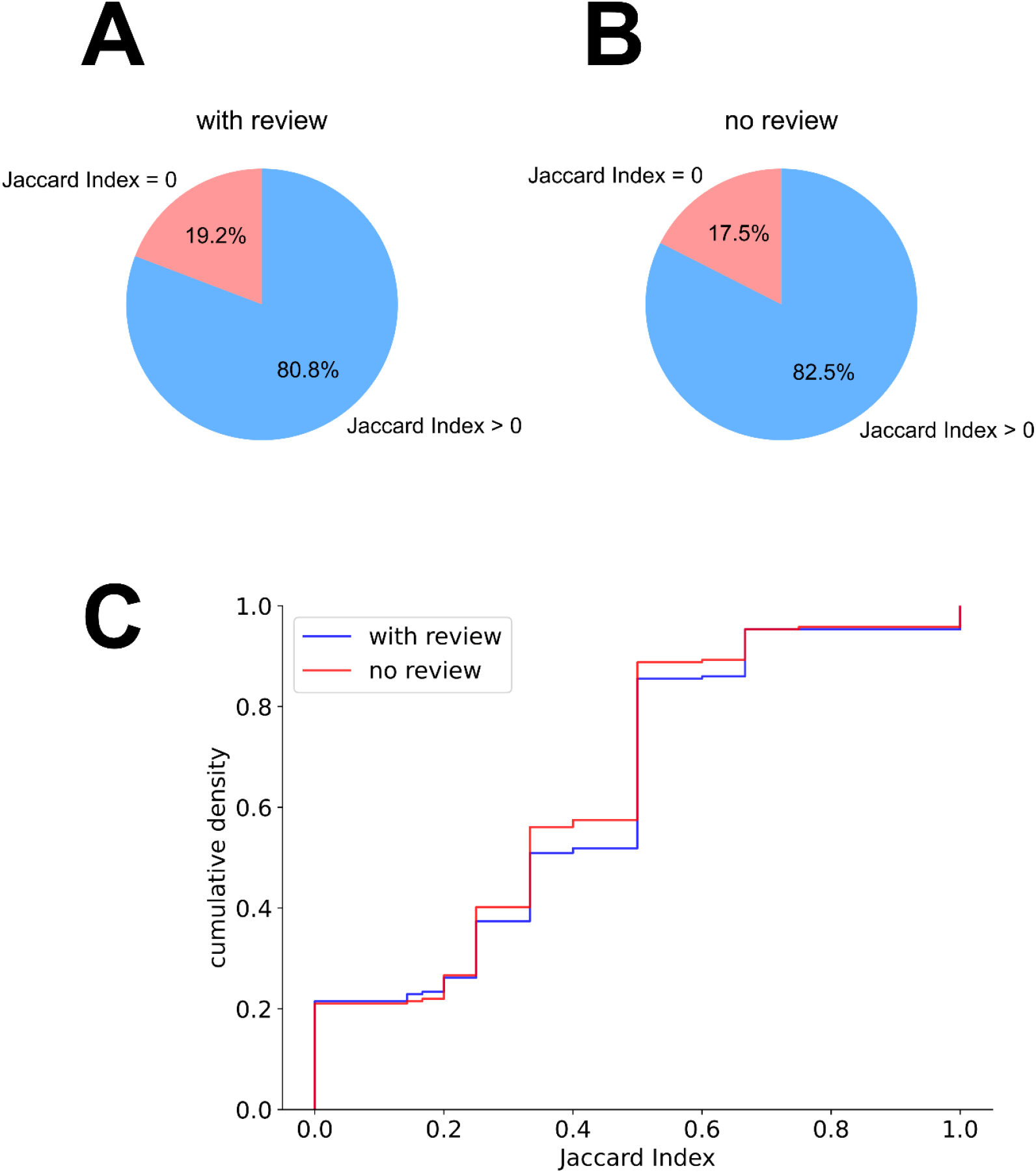
Comparison of contact selection between expert-selected patient settings and algorithm recommended settings. **(A-B)** Partial overlap in calculated contact selection with the *expert-selected* settings, **(A)** without and with **(B)** the integration of the clinical review information. **(C)** Empirical cumulative distribution of the Jaccard indices *between expert and algorithm contact selections*, when using (blue) and not using (orange) the clinical review data.

For the same current as the recorded patient settings, the VTA generated with the algorithm-based selection of contacts outperformed the manual expert settings (Figure 3A – detailed breakdown presented in Table S1). To evaluate the relative efficacy of the contact selection by the algorithm, we normalised the calculated VTA overlap with the motor-STN subregion (Figure 3B), and the portion of the calculated VTA that falls within the motor-STN boundaries (Figure 3C), by dividing with the currents recommended by the algorithm, and compared them with the corresponding motor STN and VTA fractions when using the expert-selected settings – also normalised by the current indicated by the expert settings. For both comparisons we observed an improvement in terms of the normalised overlaps over the expert settings. The difference was particularly pronounced for the VTA containment within the motor STN (Hedges’ g ≥ 1.17, p < 5e-25), and slightly smaller for the motor STN coverage (g > 0.27, p < 1e-5). The inclusion of clinical review data had only a minor influence on these measures (motor STN coverage: g = 0.13, VTA containment: g = 0.15).

**Figure 3.**
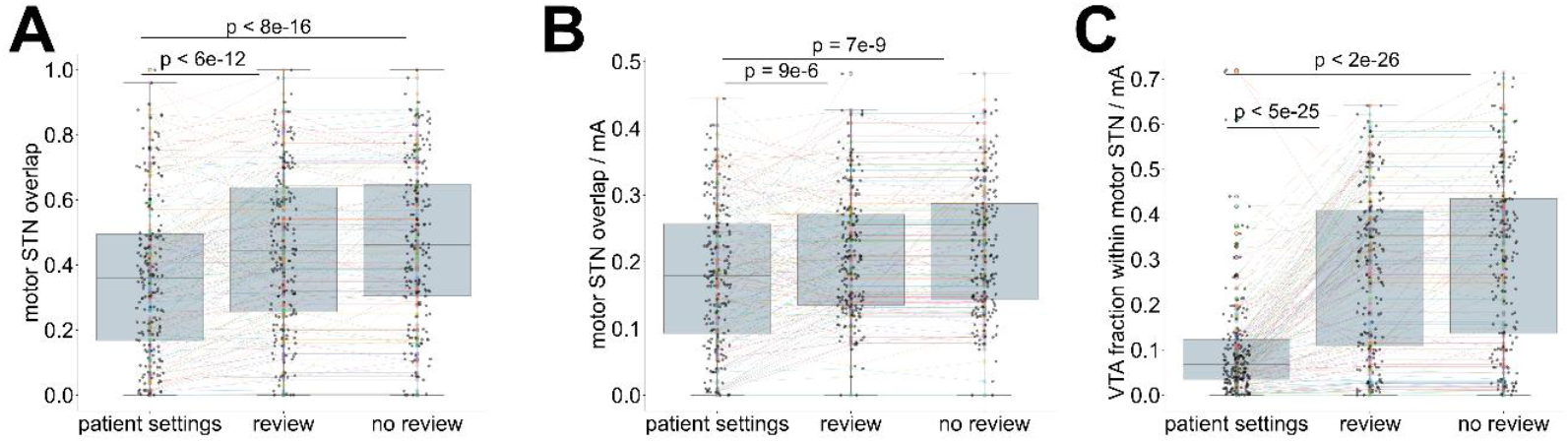
Comparison of calculated settings (with or without review information) to the *expert-selected* patient settings. **(A)** coverage of the motor STN *with* the *expert-selected current from the patient* settings, *using algorithm-suggested contact selection (with and without clinical review integration) and expert-selected contacts (patient settings)*, **(B)** current-normalised coverage of the motor STN *using algorithm-suggested settings (contacts and currents – with and without clinical review integration) and expert-selected settings (patient settings)*, **(C)** current-normalised VTA contained within the motor STN *using algorithm-suggested settings (contacts and currents – with and without clinical review integration) and expert-selected settings (patient settings)*. Wilcoxon test p-values are indicated for paired comparisons.

### Predicted improvement

To evaluate the potential benefit of the calculated settings on patient outcomes, we compiled a subset of 50 patients with a recorded DBS evaluation using UPDRS-III in the first three years after surgery. An improvement ratio was calculated from the difference of UPDRS-III scores with DBS stimulation on vs DBS stimulation off, normalised by the DBS-off score.

To estimate the expected improvement associated with the calculated settings, we trained a random forest regressor using two predictors: current-normalised motor STN coverage (reflecting contact selection effectiveness), and the stimulation current applied.

The regressor predicted the observed improvements with MSE = 0.0043, R^2^ = 0.86, indicating an acceptable fit. Predicted improvement scores were comparable between clinical and algorithm-derived settings, with a modest but statistically significant advantage favouring the calculated parameters.

This effect was consistent regardless of whether clinical review data were incorporated, with Hedges’ *g* ranging from 0.21 to 0.27 and Wilcoxon *p*-values between 0.045 and 0.019. (Figure S1)

## Discussion

The method developed here aims to provide a faster and accurate alternative to the trial-and-error approach of manual DBS programming. Following existing guidelines and practices [2], we prioritise monopolar stimulation settings. While the selected contacts could also be employed in bipolar configurations—potentially enhancing electric field steering and expanding target coverage—this would require additional considerations regarding contact polarity. Accurate polarity assignment typically depends on detailed neuroanatomical (through tractography, at minimum) or electrophysiological data, or necessitates further clinical evaluation, which falls outside the scope of this automated approach with minimal data inputs.

### Increased contact selection accuracy

Contact selection performance was evaluated by normalising the overlap between the calculated VTA and the motor STN by the stimulation current, yielding a measure of motor STN coverage per mA. The algorithm-derived settings demonstrated a small but statistically significant increase in current-normalised motor STN activation compared to clinically selected contacts. This suggests that the proposed method identifies more effective contact configurations—at a given implant position—for selectively targeting the motor subregion of the STN.

Similarly, we calculated the motor-STN and VTA overlap as a fraction of the VTA, also normalised with the current. Comparing this variable between manually selected clinical settings and the calculated settings showed that a larger portion of the VTA (per current unit) is contained within the target region (motor STN) when using the calculated settings compared to the manually adjusted settings. This significantly reduces electric field leakage in the milieu, by containing more of the high-intensity area within the target region.

Taken together, the differences in these two variables show better selective activation of the target area (motor STN in this case) when using the calculated contact selection.

### Discrepancies with the clinical settings

Exact matches occurred in only nine implantation instances without clinical review input and in ten instances when clinical review data were included, due to the procedural difference in contact selection between the two methods. The algorithm, with the settings used in this study, selects up to three sub-contacts out of a maximum of eight (in the case of directional electrodes). The clinicians, however, predominantly select entire (sub-)contact groups, matching the clinical review resolution. Thus, the only perfectly matching cases are limited to non-directional electrodes where up to two (out of four) contacts are selected by both methods.

The algorithm’s current selection strategy aims to scale the VTA to reach the midpoint of the target structure, optimising stimulation precision. Although this approach yields lower absolute coverage compared to manually adjusted clinical settings, current-normalised analyses reveal greater efficiency per unit of current. To mitigate potential under-estimation of the current with the automated method and allow further fine-tuning we provide an additional tool to facilitate exploration of different currents and their effect on STN subregion coverage.

### Comparison with alternative methods

Most recently proposed methods for initial DBS settings optimisation aim to improve the patient outcome directly [4–6] or approximate a desired volume of tissue activation [18], using statistics from large patient data pools. On the contrary, the method we described here, aims to optimise the stimulation of brain structures (STN in this instance) modulating the symptoms of the disease.Among data-driven methods, our approach most closely aligns with the StimFit toolbox [6], which also leverages patient-specific MRI data for automated DBS programming.

Our small predicted improvement, based on retrospective analysis, is also comparable with the non-inferiority to standard care achieved by the StimFit solution [19]. However, our method introduces several key advantages: it integrates clinical review data when available, explicitly targets the motor subregion of the STN to optimise motor symptom control and operates without the need for pre-trained statistical models—substantially reducing computational and data acquisition demands.

Other state-of-the-art approaches, such as Bayesian optimisation and particle swarm optimisation, have been proposed to efficiently explore the stimulation parameter space. Bayesian optimisation has been applied to reduce the number of required trials by iteratively minimising rigidity based on prior clinical responses [20], while particle swarm optimisation method was designed to address complex electrode configurations with segmented leads, and optimise stimulation amplitudes. [8]

However, these techniques are primarily outcome-driven and do not currently provide structural substrate-centric metrics or predicted improvement values, limiting the feasibility of direct comparison with our anatomy-based method.

### Limitations

The method relies on pre-operative and post-operative MRI data. Since the brain structures are directly derived from the pre-operative images and the electrode position is derived from the post-operative scan, the output is sensitive to the time delay between pre-and post-operative inputs. Increased time intervals between the pre-operative MRI scans and the post-operative scans may introduce errors both in electrode localisation relative to the target structures, due to random electrode drifts, as well as changes in the localisation of the brain structures themselves, as a result of aging or disease progression. This is expected to progressively reduce the accuracy of the method for long-term parameter adjustments, without error corrections for the MRI inputs.

Our method is considering two parameters: contact selection, and current selection. Multiple additional parameters have to be assigned by the clinicians, including frequency of stimulation, pulse width/shape, possible interleaved stimulations, and bipolar settings. Most of these parameters are usually preset to well-documented values, and are only changed later in the decision process, if necessary. [2] Accordingly, we prioritised the most time-intensive component of initial DBS programming: contact and current selection.

In the case of current estimation, our method targets the centroid of the motor STN subregion while applying predefined lower and upper bounds to ensure clinical plausibility. Thus, the suggested current values should be interpreted as an informed starting point rather than definitive therapeutic settings.

Additionally, in the present study, the input of the clinical review data was weighed equally with the geometry-based calculation. This may be unsuitable if the clinical review information is highly prioritised, for instance, to ensure a minimum stimulation effectiveness or minimising side effects. In such scenarios, increasing the weighting of clinical inputs may enhance the method’s alignment with clinical priorities and patient-specific therapeutic goals.

Finally, this study was retrospective in nature, with predicted motor improvement derived from a subset of patients who had early UPDRS-III evaluation records following DBS implantation. Although the predicted outcomes are consistent with those reported by comparable similar methods, a prospective, controlled clinical trial will be necessary to validate the effectiveness of this approach with greater certainty.

### Conclusion

This study presents a personalised, interpretable method for DBS parameter selection that leverages routinely acquired anatomical imaging and optionally available clinical evaluation data.

By integrating patient-specific MRI with clinical evaluation data and established DBS modelling tools, we demonstrate a significant improvement in targeting a motor improvement-related structure, and predict a small, but statistically significant improvement of clinical outcomes. This represents a promising patient-specific, low-overhead improvement over common clinical standard practice, aiming to accelerate the initial DBS optimisation steps close to their optimal state for targeting PD motor symptoms.

## Data Availability

All data produced in the present study are available upon reasonable request to the authors

## Author Contributions

Conceptualization: P.F., D.N., Data curation: A.L., P.F., Formal analysis: A.M., Funding acquisition: D.N., Investigation: A.M., A.L., Methodology: A.M., A.L., P.F., E.B., D.N., Project administration: D.N., Resources: P.F., D.N., Software: A.M., Supervision: D.N., Visualization: A.M., Writing – original draft: A.M., Writing – review & editing: A.M., A.L., P.F., E.B., D.N.

## Funding

The study was supported by the Brain Dynamics, grant number, CZ.02.01.01/00/22_008/0004643.

### Acknowledgments

We would like to thank the iTEMPO office (Neurology clinic, General University Hospital in Prague) for the provision of patient information.

## Data and code availability

The data is available upon request.

The code is available at https://doi.org/10.5281/zenodo.15737896 under the terms of the GNU General Public License version 3.

