## Supplementary Material for "Patient-Specific and Interpretable Deep Brain Stimulation Optimisation Using MRI and Clinical Review Data"

Table S 1. Paired Wilcoxon tests between patient settings, algorithmically calculated settings with and without clinical review data.

| **motor STN overlap (patient settings current)** | | |  |
| --- | --- | --- | --- |
|  | **No review** | **Review** | **Patient settings** |
| **No review** | Median=0.462, N=177 | p = 0.039, g = 0.085 | p = 7.863e-16, g = 0.426 |
| **Review** | - | Median=0.445, N=177 | p = 5.045e-12, g = 0.342 |
| **Patient settings** | - | - | Median=0.361, N=177 |
| **motor STN overlap (initial centroid estimate)** | | |  |
|  | **No review** | **Review** | **Patient settings** |
| **No review** | Median=0.251, N=177 | p = 0.105, g = 0.119 | p = 2.150e-07, g = 0.611 |
| **Review** | - | Median=0.233, N=177 | p = 1.104e-08, g = 0.667 |
| **Patient settings** | - | - | Median=0.361, N=177 |
| **VTA fraction within the motor STN** | | |  |
|  | **No review** | **Review** | **Patient settings** |
| **No review** | Median=0.324, N=177 | p = 4.309e-04, g = 0.145 | p = 3.884e-27, g = 1.121 |
| **Review** | - | Median=0.297, N=177 | p = 4.133e-25, g = 0.995 |
| **Patient settings** | - | - | Median=0.141, N=177 |
| **current-normalised motor STN overlap** | | |  |
|  | **No review** | **Review** | **Patient settings** |
| **No review** | Median=0.208, N=177 | p = 1.197e-02, g = 0.133 | p = 7.137e-09, g = 0.399 |
| **Review** | - | Median=0.203, N=177 | p = 8.957e-06, g = 0.279 |
| **Patient settings** | - | - | Median=0.179, N=177 |
| **current-normalised VTA fraction within the motor STN** | | |  |
|  | **No review** | **Review** | **Patient settings** |
| **No review** | Median=0.324, N=177 | p = 3.022e-04, g = 0.146 | p = 1.338e-26, g = 1.293 |
| **Review** | - | Median=0.297, N=177 | p = 4.836e-25, g = 1.167 |
| **Patient settings** | - | - | Median=0.068, N=177 |
| **current (mA)** | |  |  |
|  | **No review** | **Review** | **Patient settings** |
| **No review** | Median=1.0, N=177 | p = 1.074e-01, g = 0.041 | p = 2.132e-16, g = 0.888 |
| **Review** | - | Median=1.0, N=177 | p = 1.329e-15, g = 0.848 |
| **Patient settings** | - | - | Median=2.0, N=177 |

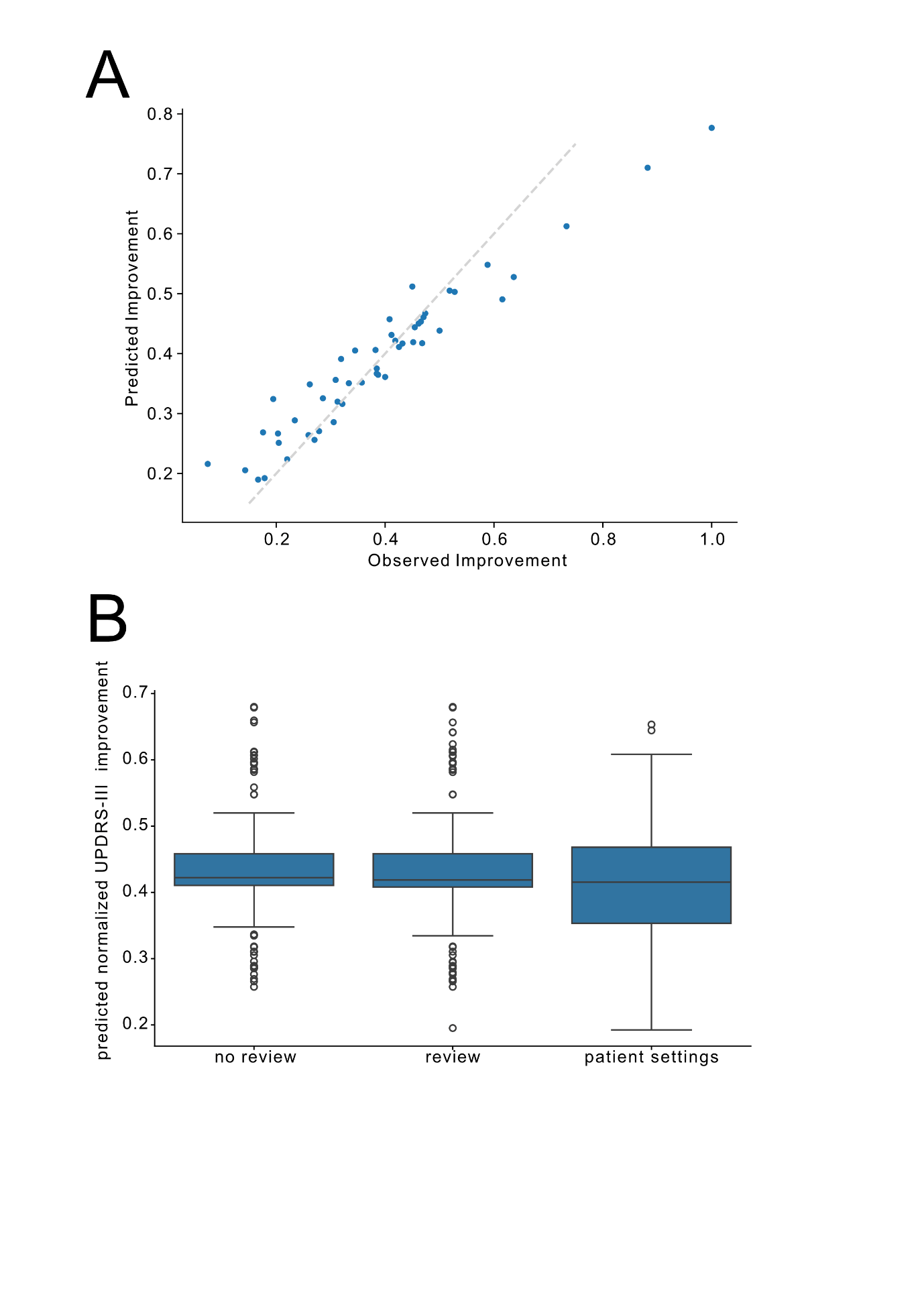

Figure S 1. Non-linear model prediction of UPDRS-III improvements. (A) Observed-Predicted plot for the 50 patients with DBS settings and UPDRS evaluations within 3 years post-implant, using a Random Forest model (5000 trees, maximum depth 7 splits, R^2^ = 0.86, MSE = 0.004). The 45-degree line is shown in dashed grey. (B) Prediction using the model on the settings derived by the algorithm (without and with integration of the clinical review information – first two box plots) and the settings assigned manually (third boxplot).

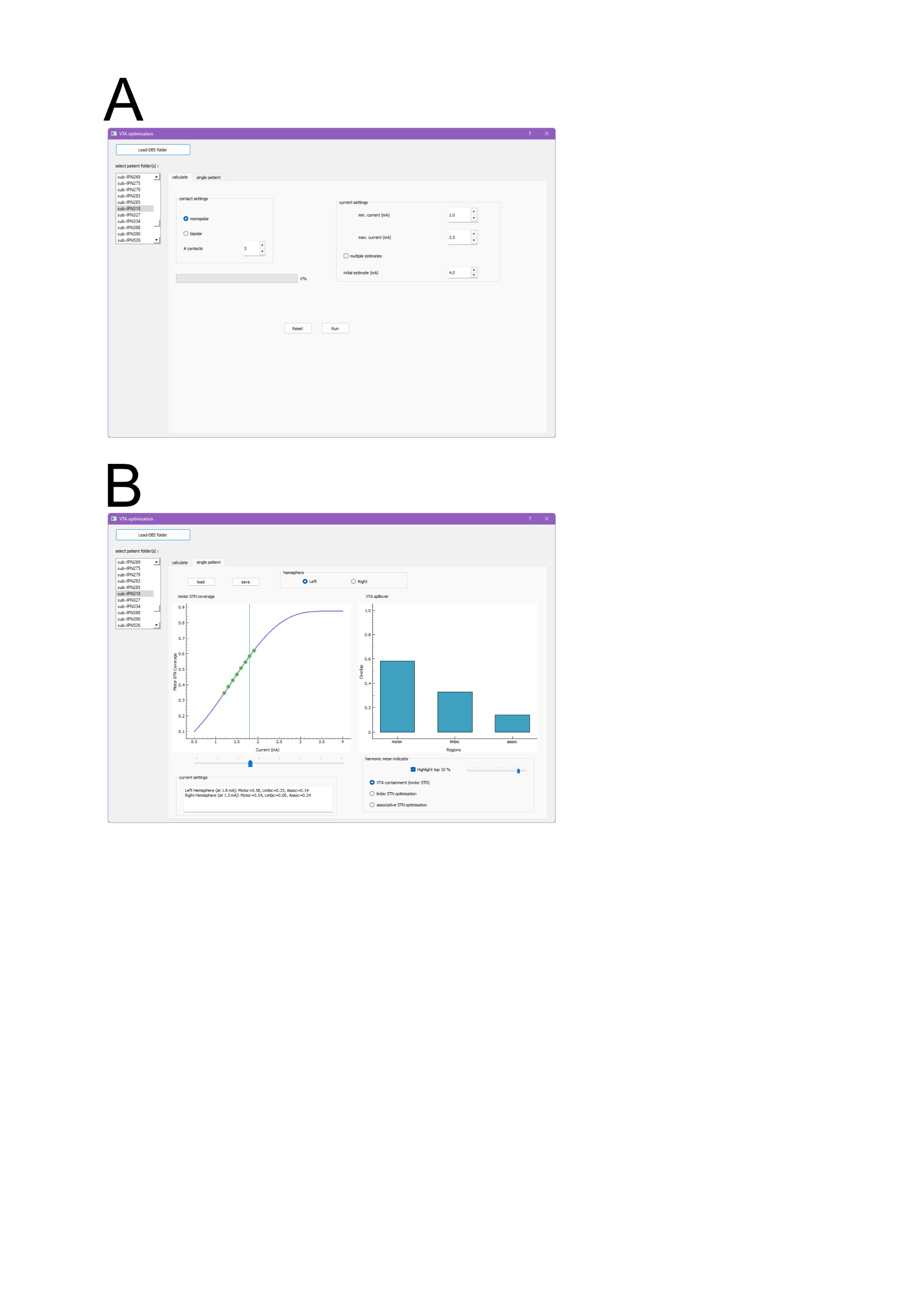

Figure S 2. User interface. (A) The first page includes a minimal number of options to guide the automated contact and current suggestions. This includes a maximum number of contacts to be considered, an option to calculate the VTAs assuming bipolar stimulation, current limits and initial value, as well as an option to calculate VTAs for multiple (n=4) currents for increased interpolation accuracy. (B) The second page allows user-guided current optimisation based on the data generated in the calculation step. A plot of the main target overlap (motor STN) vs current is shown on the left panel, using a cubic spline fit. Additional STN subregion overlaps (VTA leakage) are shown in the right panel for the selected current (using the slider). All values (for both left and right hemispheres) are updated in the “current settings” information box. A highlighter function calculates the harmonic means for every possible current (using the cubic spline fit data) between motor overlap and either the VTA containment within the motor STN or the non-overlapping part of the limbic/associative STN subregions. A small slider adjusts the marked points (green dots on the left panel plot) to reflect a user-selected percentage from the top of the selected harmonic mean range. A “save” button exports the complete settings for the current patient folder (for both hemispheres) including the contact and user-adjusted current selections as a printable HTML page.
